# A Systematic Review of the Attitude, Beliefs, and Acceptance of COVID-19 Vaccine in the Western and Eastern Hemisphere

**DOI:** 10.1101/2023.11.08.23298274

**Authors:** Roopa Kumari, Sindhu Luhana, Om Parkash, FNU Sadarat, Subhash Chander

## Abstract

**Background:** The availability of an effective vaccine does not equate to its use; its effectiveness primarily depends on vaccine acceptance by the targeted population. Despite the rapid development and widespread access to the COVID-19 vaccine, herd immunity is yet to be achieved, with vaccine hesitancy as a major barrier. This study sought to systematically assess the beliefs, attitudes, and acceptance towards COVID-19 vaccines, including factors contributing to vaccination hesitancy in the Eastern and Western hemispheres.

**Methods:** A comprehensive search of articles was conducted through Scopus, PubMed, Embase, CINAHL, Cochrane CENTRAL, and Web of Science databases for studies published from inception to May 2023 using the PRISMA guidelines.

**Results:** Our search yielded 1154, of which 21 were eligible for inclusion. The rate of willingness or intention to vaccinate varied with the geographic region, from 12% in the USA to 93.9% in China. Four studies from the Western and two from Eastern regions reported a low acceptance rate (defined as <50%): USA (12%), Spain (48.3%), Switzerland (38.6%), Europe (multi-national, 31%), Nepal (38.3%), and Oman (43%). Overall, vaccine acceptance was low-to-moderate in the general population and healthcare workers (HCWs) in both Eastern and Western hemispheres except for China which reported high acceptance (defined as >75%) among the general population and HCWs. Demographic characteristics (female, younger age, and higher education) and non-demographic factors (knowledge about the COVID-19 vaccine and its development, history of influenza vaccination, perceived susceptibility or severity of infection, and the belief that vaccines are effective in controlling the pandemic were associate with high acceptance rates or intentions to take the COVID-19 vaccine. On the other hand, mistrust of the vaccine, its safety and effectiveness, disinformation or poor awareness of the vaccine, side effects concerns, belief in natural immunity, previous adverse experience with the vaccines, and distrust in the information sources about the COVID-19 pandemic were associated with vaccination hesitancy.

**Conclusion:** For better acceptance, COVID-19 vaccination campaign strategies should be modeled based on regional political, economic, and social contexts.

**Highlights:**

- The willingness to accept COVID-19 vaccines was low-to-moderate in both hemispheres.
- Healthcare workers were as willing to vaccinate as the general population.
- Vaccine hesitancy was associated with demographic and vaccine-specific factors.

## Introduction

The COVID-19 pandemic is one of the most significant pandemics of the 21^st^ century accounting for over 767 million infections and nearly 7 million deaths as of July 2023 [1]. Due to its severity, pandemic control measures such as social distancing were prioritized alongside developing novel vaccines and pharmaceutic and biologic agents to treat patients with severe symptoms [2]. There is a global consensus that vaccination is the imminent and ultimate key to controlling the COVID-19 pandemic. For instance, in May 2020, the 73^rd^ World Health Assembly issued a resolution that attributed comprehensive immunization as a global fundamental public health strategy to prevent, contain and stop the spread of COVID-19 [3].

In response to the call, multiple COVID-19 vaccines were developed and tested across diverse populations, signifying a significant breakthrough in vaccine development to contain the pandemic. The US Food and Drug Administration issued the first emergency use authorization for the BTN162b2 vaccine (Pfizer-BioNTech) on 11 December 2020 to inoculate young adults aged 16 years and above [4]. Soon after, emergency use authorization was granted for mRNA-1273 (Moderna), AZD1222 (Oxford/AstraZeneca), and Ad26.COV2.5 (Janssen) vaccines [5]. By the first quarter of 2021, there were 85 vaccine candidates in the clinical trial phase and 184 in the pre-clinical phase [6].

However, reluctance to accept a potential vaccine was reported even before the widespread availability of COVID-19 vaccines in the general population and healthcare workers (HCWs) [7, 8]. COVID-19 vaccine hesitancy has been a barrier to scaling up the vaccination rate for achieving herd immunity [9]. Vaccine hesitancy is a multi-factored phenomenon rooted in several factors such as an individual’s socio-political stand, religious convictions, trust in government or the healthcare system, health/vaccine literacy, the source of information, and the perceived severity or risk of contracting the disease [10]. These factors tend to aggregate with other vaccine-specific factors, and as a result, the underlying cluster of factors of vaccine hesitancy varies by the vaccine. In the case of COVID-19, the rapid development and regulatory approval of the vaccine, political affiliations, and widespread misinformation about vaccine safety, efficacy, and disease severity have played a key role in low public confidence in the vaccine [11, 12]. As a result, significant disparities in regional vaccination rates have emerged. As of 26 June 2023, persons vaccinated with a complete primary series per 100 population stood at 31.5 for Africa, 51.3 for Eastern Mediterranean, 64.6 for Europe, 69.2 for South-East Asia, 71.2 for the Americas, and 85.4 for Western Pacific regions [1].

Given the disparities in vaccination rates, region-specific interventions may be required to improve the willingness to vaccinate. Although several studies have explored the factors associated with COVID-19 at the country level, regional studies are scarce. Therefore, the present study sought to conduct a systematic review of the current works of literature regarding the facets of vaccine hesitancy, particularly the factors associated with the beliefs, attitudes, and acceptance of vaccines in the Western and Eastern hemispheres.

## Methodology

A rapid systematic review approach was utilized in conformity with the guidelines of the Preferred Reporting Items for Systematic Reviews and Meta-Analysis (PRISMA) reporting statement [13] to give valid and timely evidence that informs various decision-making concerning COVID-19 vaccination strategies.

### Search strategy

A systematic literature search was conducted for relevant articles published from inception to May 2023 in the following databases with customized retrieval strategies for each database: Scopus (Medline), PubMed, Embase, CINAHL, Cochrane CENTRAL, and Web of Science. Regarding the search syntax, the controlled vocabulary involved generic free-text search terms such as ‘population,’ ‘willingness,’ ‘attitude,’ ‘beliefs,’ and ‘vaccine hesitancy.’ The MeSH vocabulary terms also included ‘vaccine,’ ‘COVID-19 vaccines,’ ‘sars-cov-2 vaccine,’ ‘vaccination refusal,’ ‘2019 novel coronavirus,’ and ‘coronavirus.’ The English synonyms were also used systematically as search syntax items in the databases to minimize the chances of missing relevant studies. In addition, the lists of bibliographies of the eligible articles and Google Scholar were manually searched to identify any potential reference for inclusion.

### Inclusion and exclusion criteria

The eligibility of the articles obtained from the database and bibliography searches was assessed using the following inclusion criteria:

i. Studies involving the general public and/or HCWs. HCW was defined as persons employed or studying within the healthcare facility, including physicians, medical studies, and the ancillary staff.
ii. Published primary studies inclusive of structured design, such as cross-sectional studies and semi-structured surveys.
iii. Articles that examined the attitudes, acceptance, and or beliefs towards COVID-19 vaccination and any related factors. In this accord, vaccination acceptance was defined as the percentage of the population willing to accept vaccination when available.
iv. The English language articles were prioritized for inclusion, and any other relevant articles could be translated into English.

Duplicated studies, commentaries, case reports, protocols, editorials, letters, conference abstracts, retracted, and articles in preprint were excluded. Articles not available online in full or from low-quality sources were also excluded.

### Article screening and selection

All the articles from the search were imported into the Endnote software [14] and deduplicated. Titles and abstracts of the remaining articles were assessed, and those not meeting the inclusion criteria were excluded. The remaining articles were subjected to full-text screening. If more than one article utilized the same dataset, the one with the largest sample size was included.

Two reviewers (A.F. and J.L.) conducted article screening and selection. Any differences between the reviewers were resolved through discussion. However, if the two reviewers could not arrive at a consensus on excluding or including some articles, the senior reviewer (G.H.) was consulted.

### Data extraction and management

Abstraction of all the relevant data was simultaneously performed by two reviewers (A.F. and J.L.) independently using a standardized data extraction form and then exchanged for validation to prevent data inconsistency and errors. Any disagreement was resolved through discussion with the third reviewer (G.H.). Information that reflected the attitude or beliefs of the participants towards receiving COVID-19 vaccines were sought through either of the following measurements: rate of COVID-19 vaccine acceptance or rejection, rate of positive/negative response to specific vaccine uptake, rate of individuals willing to enroll in a COVID-19 vaccine trial. The other relevant significant information that was sought included: the first author and publication year, the geographical context of the participants (i.e., country/city/setting), study design, survey period, study population of interest and sample sizes, data collection methods, demographic characteristics, and motivation and barriers towards vaccine uptake (vaccine hesitancy factors).

The vaccine acceptance rate was defined as the proportion of the population who were willing to or had taken the locally available COVID-19 vaccine and was categorized as low (<50%), moderate (50-75%), and high (>75%).

### Evidence quality assessment and risk of bias

The selected articles’ methodological quality and risk of bias were assessed using the Newcastle-Ottawa (NOS) quality scale independently by two reviewers (A.F. and J.L.). Any uncertainties were resolved by consulting the third reviewer (G.H.). The NOS scale was developed to assess the quality of non-randomized studies of their design, content, and ease of use. The NOS has seven quality scoring systems under three main themes: selection, comparability, and outcome.

## Results

### Search results and study selection

Our database search returned 1,137 articles, with an additional 17 from manual reference list screening. Seven hundred seventy-eight articles were removed during de-duplication. Titles and abstracts of the remaining 367 studies were screened, of which 103 articles were eligible for full-text screening. Finally, 21 articles fully meeting the inclusion criteria were identified and included in this systematic review. The flow of study screening and selection is shown in **Figure 1**.

**Figure 1:**
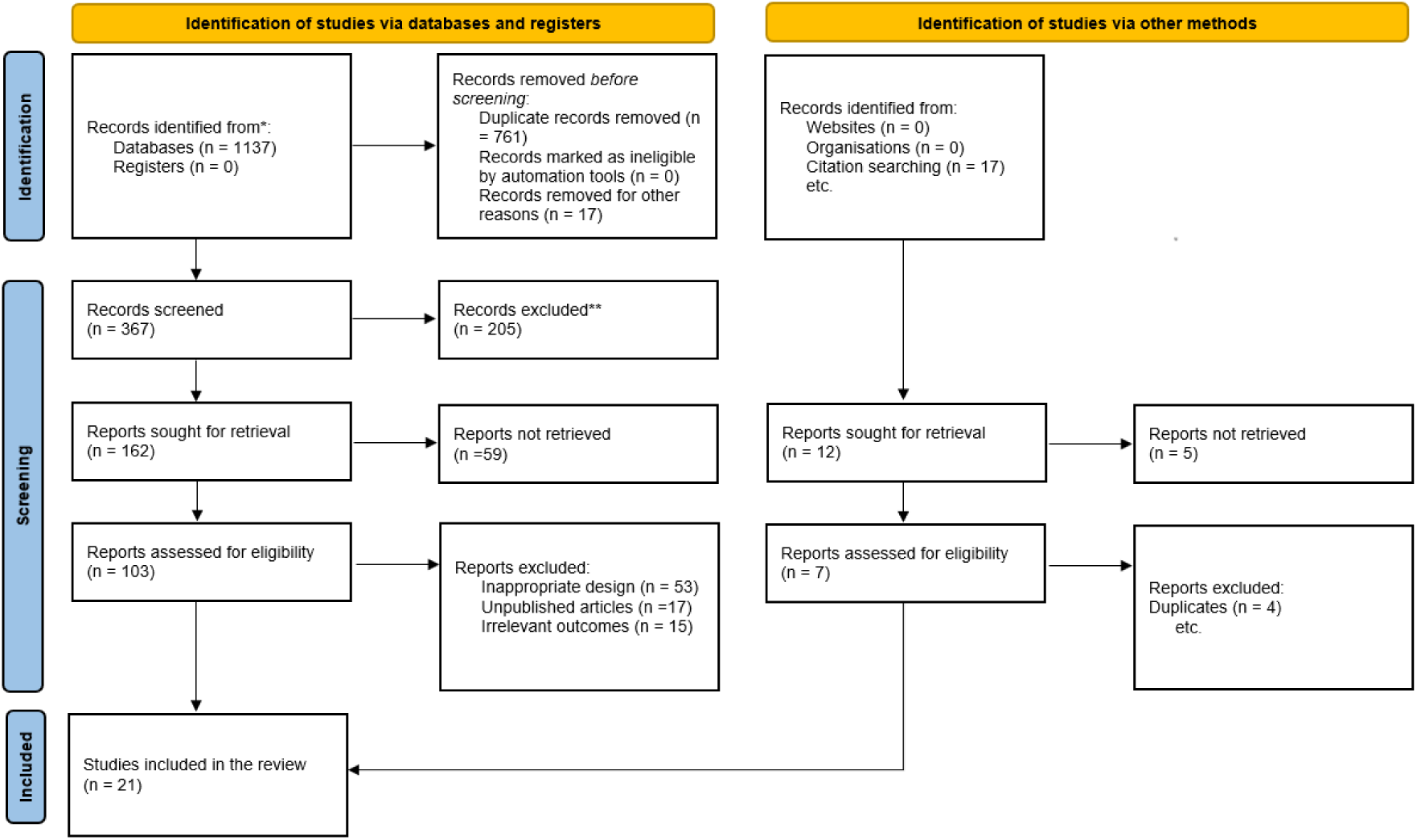
PRISMA flow chart for study screening and selection process.

### Study characteristics

The studies included in this review were published between 2020 and 2022, although most were published in 2021 (n=16). All the included studies adopted a cross-sectional design and specifically targeted adults aged above 18 years. The sample sizes ranged from 102 to 12,034 subjects.

Ten of the 21 studies were from the Eastern hemisphere encompassing India [15, 16], China [17, 18], Indonesia [19], Nepal [20], Oman [21], Saudi Arabia [22, 23], and the United Arab Emirates [24]. Studies from the Western hemisphere originated from Spain [25], Switzerland [26], Poland [27], Slovakia [28], USA [29–32], Canada [33], and Ecuador [34]. There was one European multi-national study [35].

The detailed characteristics of the included studies are presented in **Table 1**.

**Table 1:**
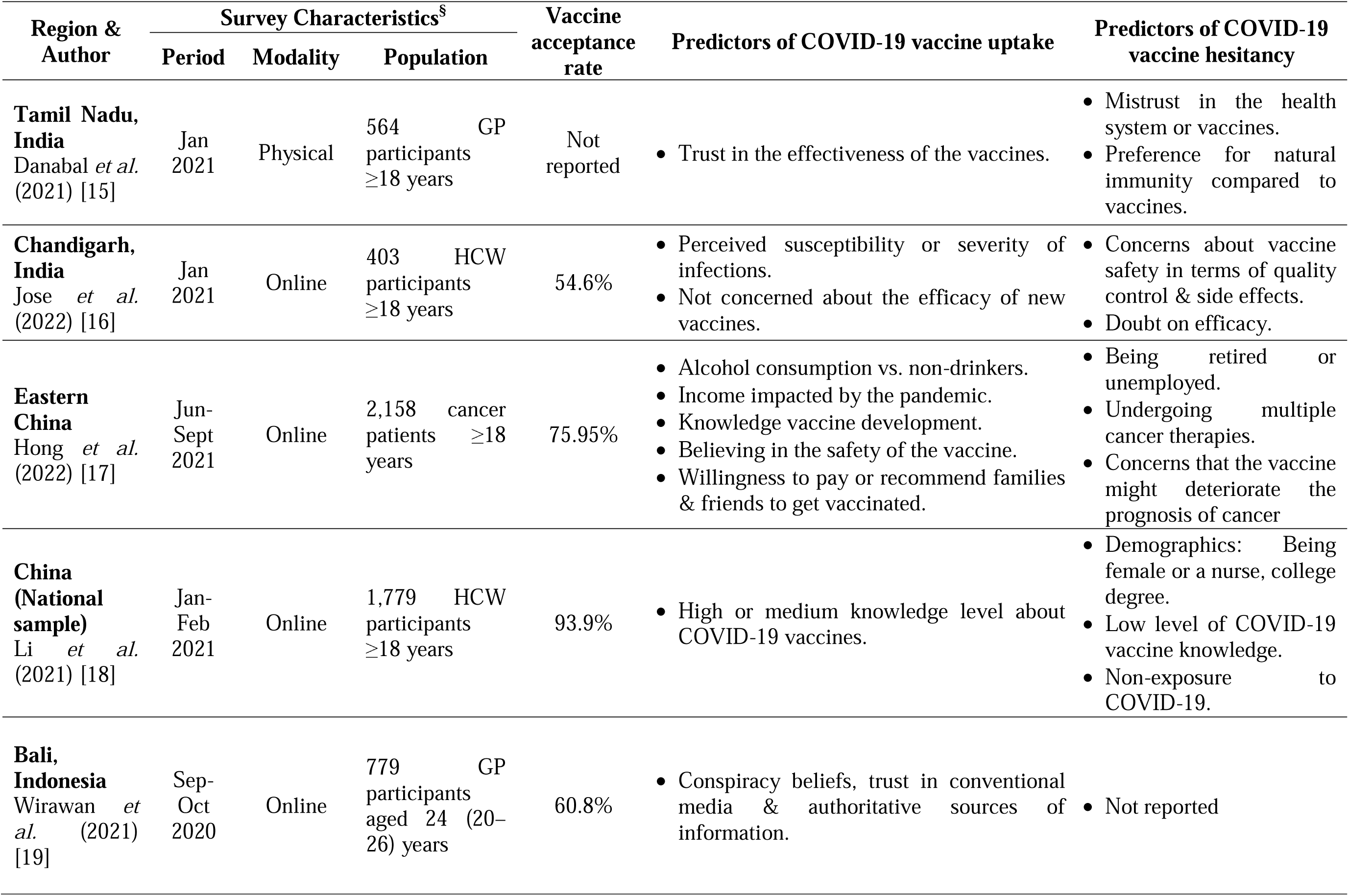
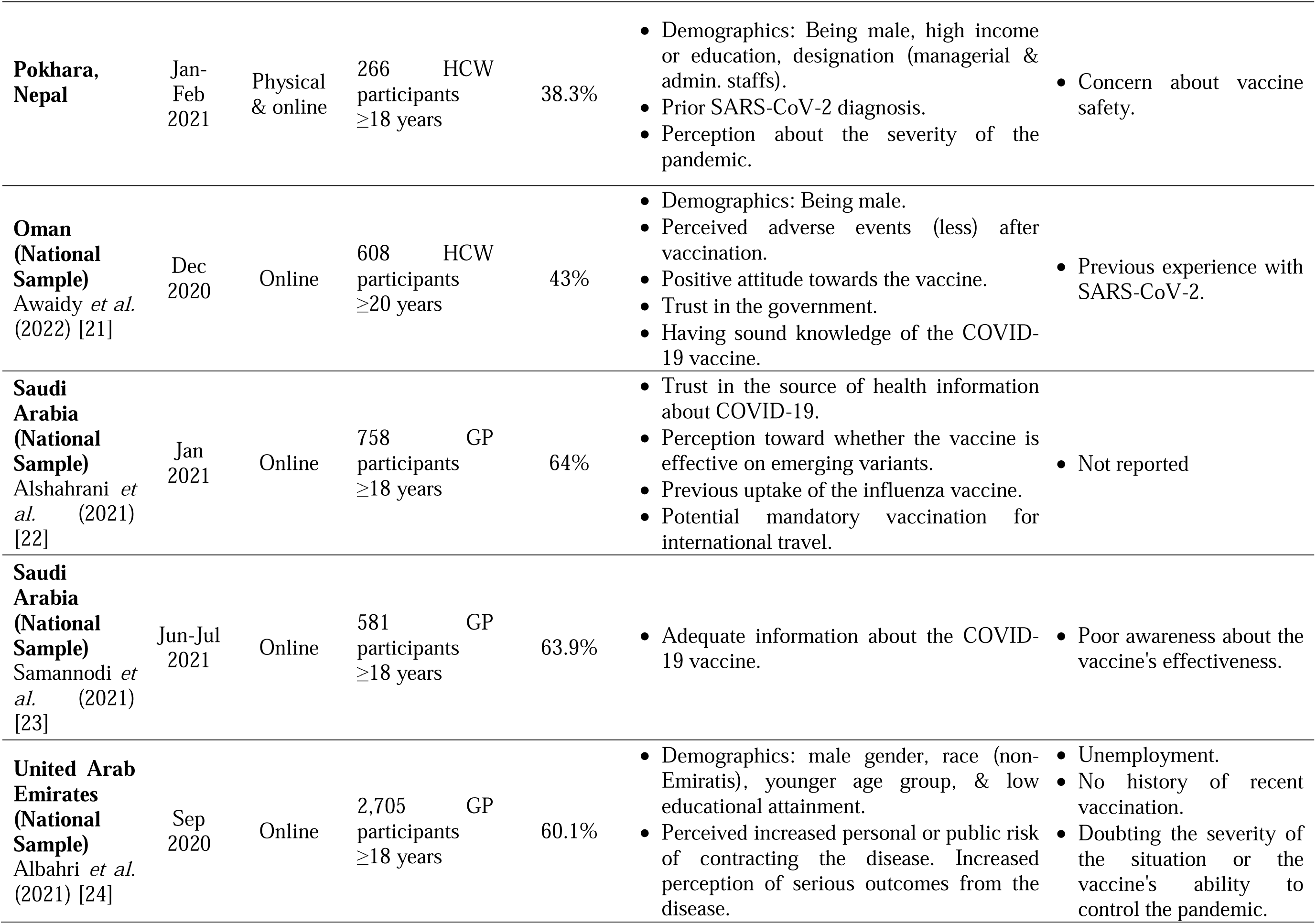
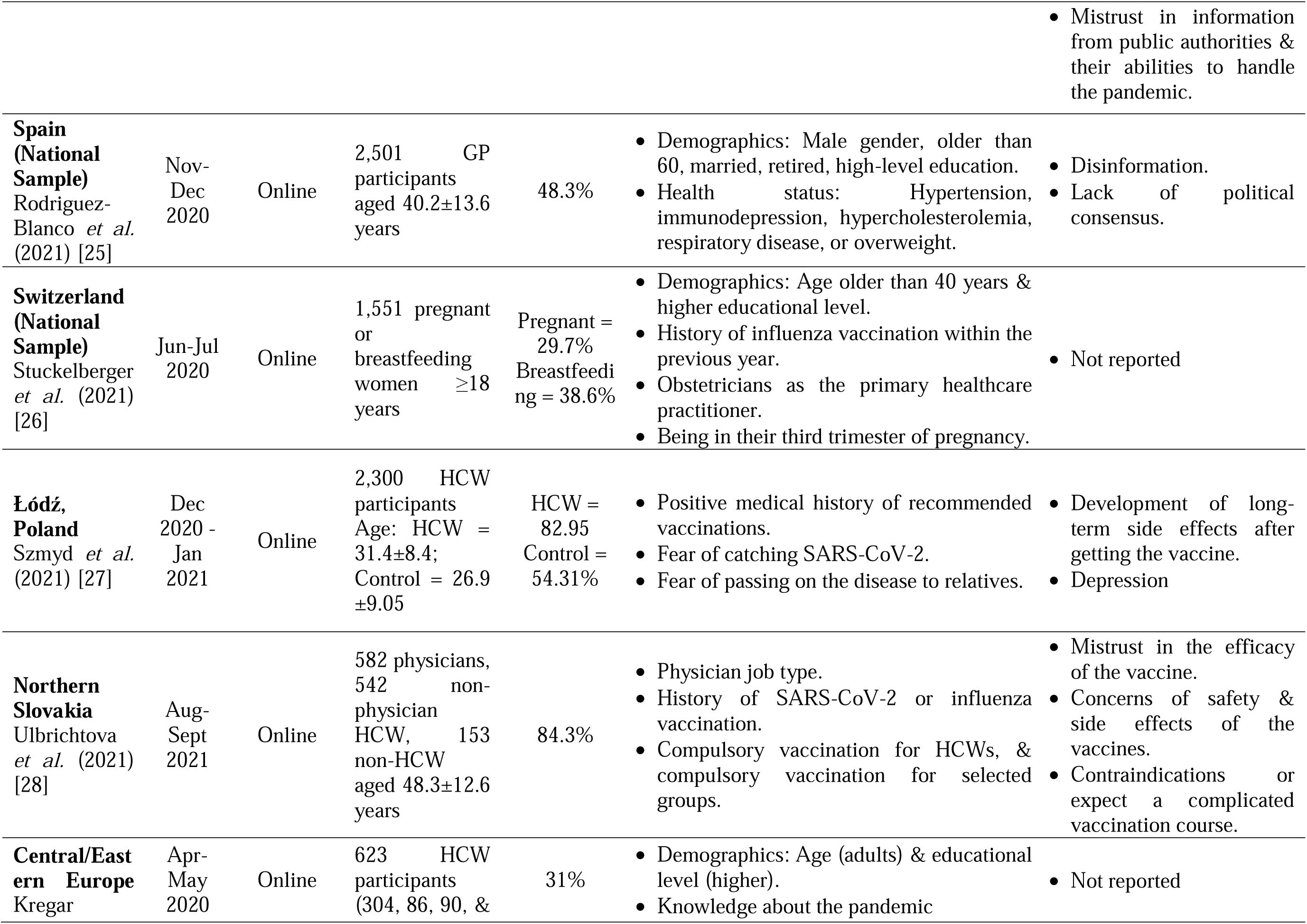
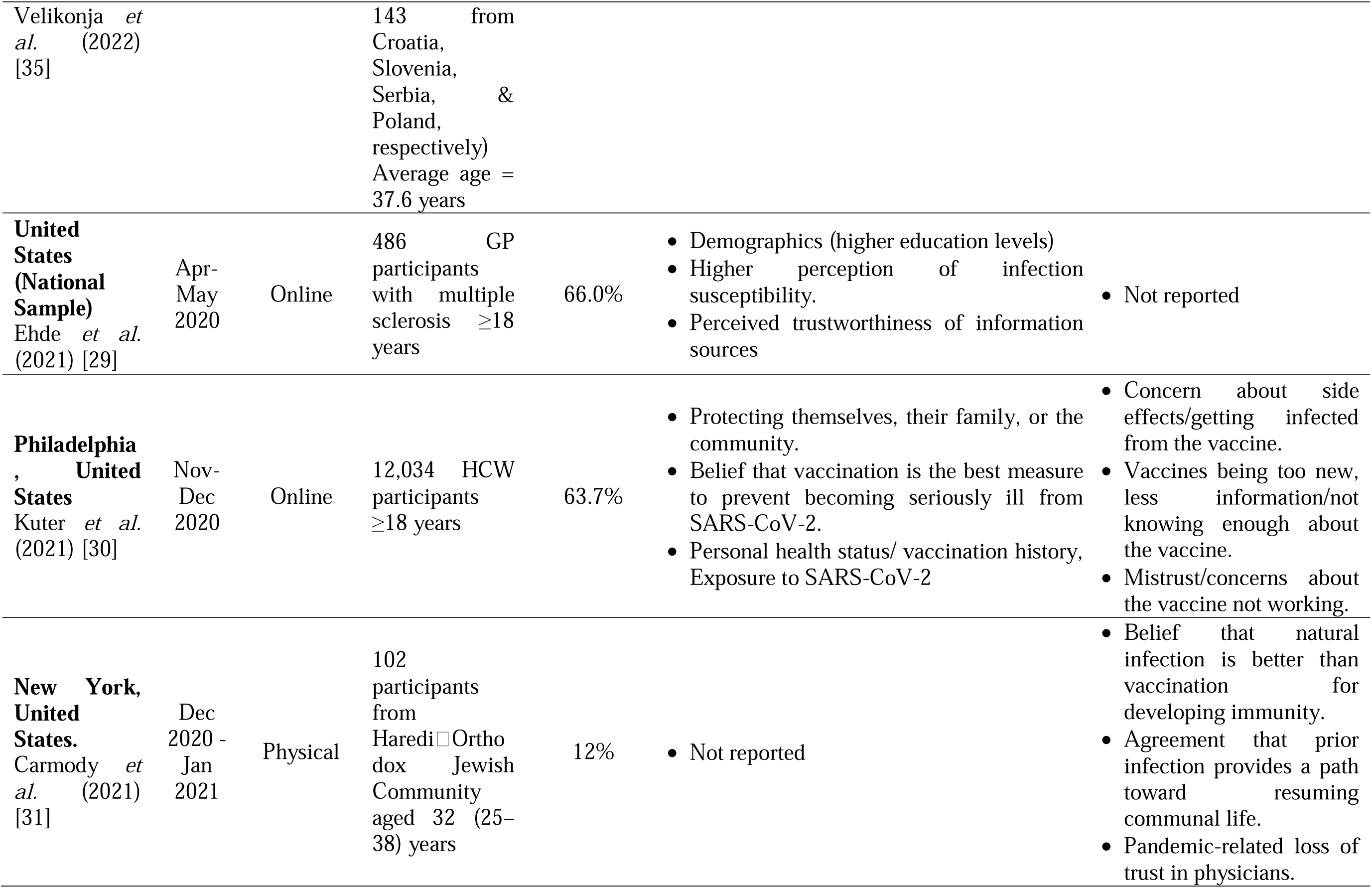
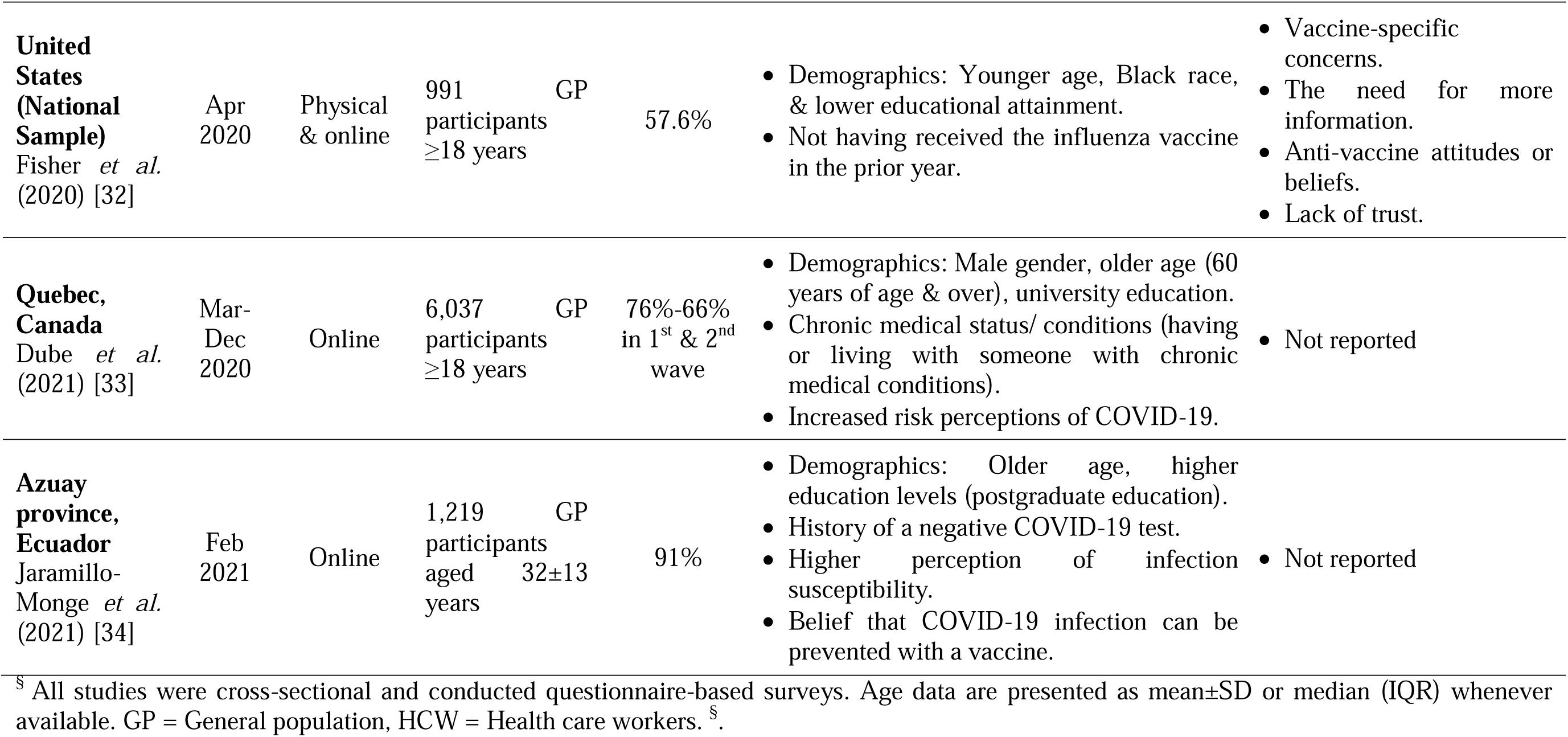
Summary of the included studies.

### Quality appraisal & assessment

The results of the quality appraisal for the included studies are presented in **Table 2**. Fifteen studies scored six, while the rest scored five on the NOS quality scale, indicating good quality.

**Table 2:**
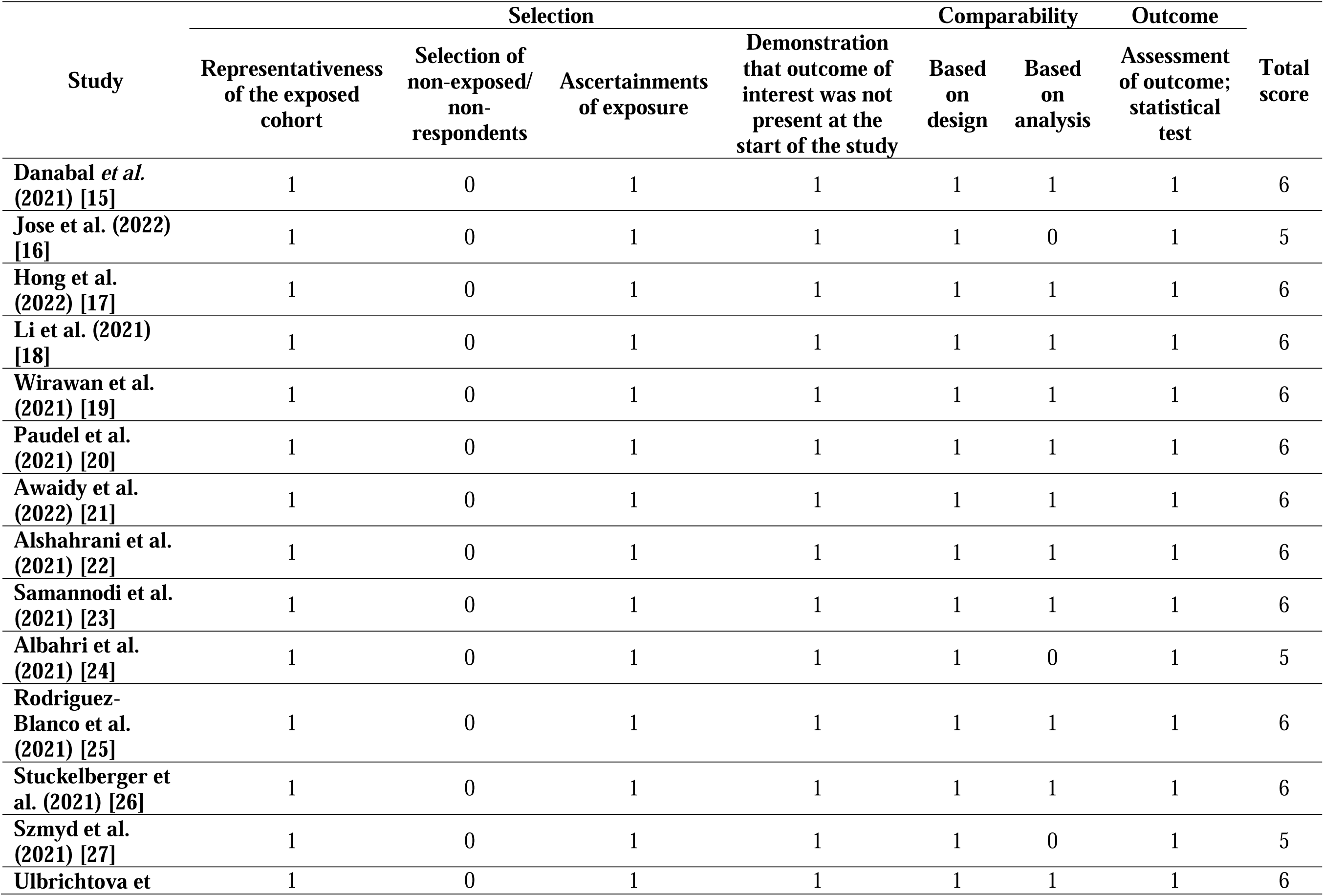
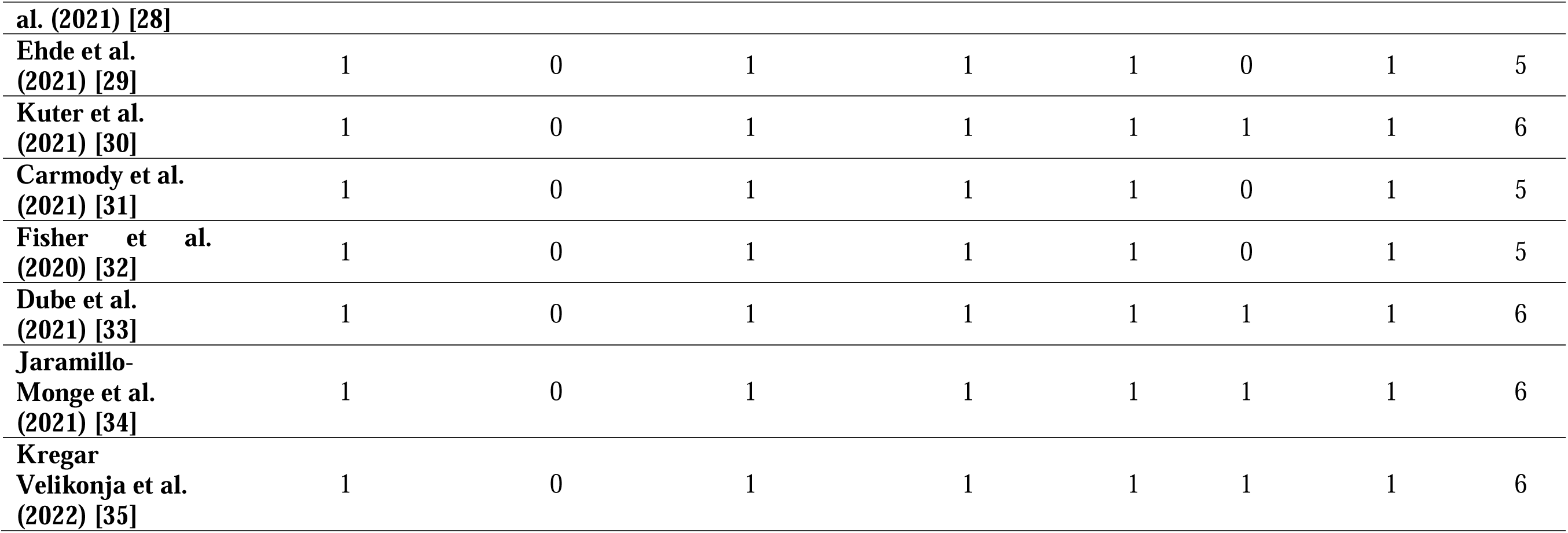
Newcastle-Ottawa scale scores for quality appraisal.

## Evidence Synthesis and Discussion

### Summary of Key Findings

A total of 21 studies were identified from 14 countries in the Eastern and Western hemispheres that explored factors associated with attitudes, beliefs, and intentions to accept the COVID-19 vaccine. Findings indicated that COVID-19 vaccine acceptance was low-to-moderate in both the general population and HCWs. Demographic characteristics, including age, gender, education, and social status, were strongly associated with the willingness to vaccinate against COVID-19. Additionally, non-demographic factors such as sound knowledge about the COVID-19 vaccine and its development, history of previous influenza vaccination, perceived risks (including fear of contracting the family, protecting self, family, or community), the trustworthiness of the information source regarding COVID-19 disease, and the beliefs that the virus can be prevented using the vaccine or a positive attitude towards the vaccine and its efficacy were associate with high acceptance rates or intentions to take the COVID-19 vaccine. On the other hand, mistrust of the vaccine, its safety and effectiveness, disinformation or poor awareness of the vaccine, side effects concerns, belief in natural immunity, previous adverse experience with the vaccines, and distrust in the information sources about the COVID-19 pandemic were associated with vaccination hesitancy.

### Acceptance rates

All but one article [15] evaluated the willingness to receive the COVID-19 vaccine. The rate of willingness or intention to vaccinate varied with the geographic region from 12% in New York, USA [31] to 93.9% in China [18]. Four studies from the Western and two from Eastern regions reported a low acceptance rate: USA (12%) [31], Spain (48.3%) [25], Switzerland (38.6%) [26], Europe (multi-national, 31%) [35], Nepal (38.3%) [20], and Oman (43%) [21].

### Demographic factors associated with attitude/beliefs/intention to accept the COVID-19 vaccine

The factors associated with the beliefs, attitudes, and intentions toward COVID-19 vaccine acceptance were analyzed in 11 of the included studies [20, 21, 24–26, 28, 29, 32–35], which used either or both multivariate and univariate logistical regression analysis to account for the confounding variables. The studies noted several demographic factors associated with COVID-19 vaccine uptake, including age, gender, education level, occupation or social status, and race. Five studies reported that the male sex strongly predicted acceptance or higher intentions to vaccinate against COVID-19 than the female gender [20, 21, 24, 25, 33]. Of the seven studies that analyzed age, five studies [20, 21, 33–35] indicated that being above 40 years strongly correlated with vaccine acceptance, while the other two studies showed that younger age [24, 32] predicted the intention to take COVID-19 vaccine among HCW and the general population. Six studies indicated that higher education levels [20, 25, 29, 33–35], especially degree attainment, strongly predicted higher intention towards COVID-19 vaccine acceptance, contrary to two studies that showed that low education attainment [24, 32] was associated with a high likelihood of vaccination acceptance. Regarding social status, studies indicated factors such as marriage, retirement, and high income as determinants of COVID-19 vaccine acceptance [25]. Nevertheless, two studies reported that identifying as White was associated with a greater intention to vaccinate than those identifying as Black, Asian, Hispanic, or others [30, 32]. Among HCWs, designation or the type of work was a strong predictor of vaccination [20, 28].

### Non-demographic factors associated with attitude/beliefs/intention to accept the COVID-19 vaccine

The included studies also reported factors unrelated to population demographics that correlated with the uptake of the COVID-19 vaccine. In particular, the high-risk perceptions towards COVID-19, including fear of contracting the disease and perception towards protecting self, relatives, family, and community was identified by eight studies as a significant contributor to acceptance and intention to take COVID-19 vaccine [16, 20, 24, 27, 29, 30, 33, 34].

Furthermore, five studies weighed the belief that COVID-19 disease is preventable through vaccination, a low risk of adverse effects, or a positive attitude towards the vaccine [15, 19, 21, 30, 34] as crucial contributors to vaccination acceptance. Four other studies identified sound knowledge of the COVID-19 pandemic and vaccine development as important factors for accepting COVID-19 vaccination [18, 21, 23, 25]. Five studies also found that previous influenza vaccination correlated to the likelihood of vaccination against coronavirus [22, 26–28, 32]. Trustworthiness of the health information source about COVID-19, including media, authorities, and the government, also emanated as a strong predictor in four included studies. The other crucial factors associated with beliefs, attitudes, or intentions to take the COVID-19 vaccine included health statuses such as chronic disease conditions, beliefs in conspiracy theories, willingness to recommend, coronavirus exposure or previous diagnosis with COVID-19, and mandatory COVID-19 vaccination orders.

### Reasons for COVID-19 vaccine hesitancy (non-acceptance)

Fifteen articles identified factors associated with COVID-19 vaccine hesitancy. Eight studies identified the fear of developing severe side effects after vaccination [16, 17, 20, 24, 27, 28, 30, 32]. Most of the individuals who were skeptical of developing side effects considered the COVID-19 vaccines unsafe and thus rejected them. Individuals also failed to accept the vaccines due to mistrust; rather, vaccination hesitancy was prompted since the targeted population showed distrust in the efficacy or effectiveness of the vaccines to treat or minimalize the Coronavirus (2019) pandemic.

Four studies highlighted disinformation, lack of or poor awareness of the COVID-19 vaccine, and its efficacy as barriers to vaccine acceptance [18, 23, 25, 30]. Another critical variable was distrust in COVID-19 information sources, which included pandemic-related loss of trust in physicians, the health system, and the authorities [15, 24, 32]. Three studies also associated the belief in natural immunity, or a belief in the perception that natural infection is better than vaccination for developing immunity, as well as the anti-vaccine beliefs and attitudes to strongly predict vaccine hesitancy [15, 31, 32]. Other factors associated with vaccine hesitancy included the vaccine being new, previous adverse effects or development of long-term side effects following COVID-19 vaccination, non-exposure to COVID-19, lacking a recent vaccination history, and doubts about the severity of the COVID-19 pandemic. One study identified demographic factors, such as female, retired, or unemployed, as factors associated with non-acceptance of the COVID-19 vaccine [18].

### Regional variations in vaccine acceptance

In the Eastern regions, vaccine acceptance varied from low in Nepal and Oman to moderate in India, Indonesia, Saudi Arabia, and the United Arab Emirates. However, two studies from China reported high vaccine acceptance rates. Consistent with our findings, a previous systematic review by Sallam (2021) [36] found that vaccine acceptance in South-East Asia was relatively high (90%) compared to Middle East countries (below 30%). Al-Jayyousi *et al.* (2021) [37] attributed the low acceptance of the COVID-19 vaccine to the widespread beliefs in conspiracy theories that negatively impacted vaccine uptake. Sallam (2021) [36] also cited low trust in the government and low education levels in Middle East countries, resulting in a low willingness to vaccinate. COVID-19 vaccine hesitancy has been reported in 6.3 to 56.2% of the population in South Asian countries, with safety and efficacy concerns as predominant factors [38].

However, it is noteworthy that the two studies from China exclusively involved HCWs [18] and cancer patients [17]. Contrary to our expectations, HCWs did not demonstrate a consistently higher acceptance rate than the general population across the region. For instance, Paudel *et al.* (2021) [20] sampled 266 Nepalese HCWs with an acceptance rate of 38.3%, Awaidy *et al.* (2022) [21] sampled 608 Omani HCWs with an acceptance rate of 43%, Jose *et al.* (2022) [16] sampled 403 India HCWs with an acceptance rate of 54.6%, while Li *et al.* (2021) [18] reported a vaccine acceptance rate of 93.9% in a nationally representative sample of Chinese HCWs. These variations may be attributed to the national mandate to vaccinate HCWs in some countries [39]. For instance, vaccination against COVID-19 has been compulsory for all people employed at hospitals in China since the first quarter of 2021 [40], while countries like India prioritized but did not mandate the vaccination of HCWs [41].

In the Western regions, low vaccination acceptance was reported from European countries [25–27]. In contrast, one study from Canada [33] and two from the United States [29, 32] reported a moderate vaccine acceptance rate among the general population. While another study from the United States reported a very low acceptance rate of 12% [31], the participants were derived from a single community (HaredilJOrthodox Jewish Community). Our search strategy yielded only one article from South America (Ecuador), which reported a high vaccine acceptance rate in the general population. Although low high vaccine acceptance rates were reported among HCWs in Europe in the first quarter of 2020, later surveys reported high vaccine acceptance rates [27, 28]. Vaccine acceptance among HCWs in the United States remained moderate in late 2020 [30], on par with the general population.

Consistent with our findings, a recent meta-analysis reported 61% vaccine acceptance among general populations and 55% among HCWs in the United States [42]. However, in the United States, low vaccine acceptance may not be solely related to vaccine hesitancy. Instead, they may be driven by the syngenetic effects of several factors such as gender (women), race (Black and Hispanic), age (younger adults), political affiliations (Republican), low education and income, and residing in rural areas [42, 43]. Similarly, moderate vaccine acceptance rates have been reported in European countries in both the general population and HCWs, with low trust/confidence in the vaccines, government and medical system, and health/vaccine literacy as the main drivers of vaccine hesitancy [44, 45]. Even in the early phase of vaccine rollout, Asian countries like China and South Korea reported higher vaccine acceptance and greater trust in government than the United States, the United Kingdom, and European countries [46, 47].

Concerns regarding the efficacy, safety, and effectiveness of the COVID-19 vaccines have been major factors for vaccine hesitancy, while perceived susceptibility and severity of COVID-19, trust in the government, and demographic factors such as female gender, younger age, higher educational attainment, and high income have been associated with willingness to vaccinate throughout the pandemic [46–54]. These vaccine refusal and acceptance predictors may be directly related to COVID-19-related disinformation. As Al-Amer et al. (2022) [39] noted, myths, rumors, and false beliefs on vaccines peddled by anti-vaccine individuals and media, especially social media, threatened vaccination uptake. Similarly, Jin *et al.* (2021) [55] supported that the beliefs in conspiracy theories circulated in the media by vaccine-averse individuals regarding the vaccine’s side effects significantly influenced people’s decision to vaccinate. For instance, Reno *et al.* (2021) [56] showed that males were less likely to believe in conspiracy theories and perceived greater severity of COVID-19 disease and had a higher willingness to vaccinate, while Troiano *et al.* (2021) [57] demonstrated greater concern towards the safety of the vaccines and distrusted the quality and impartiality of the vaccine information among females with a negative effect on vaccine acceptance. Moreover, vaccine hesitancy could also result from underestimating the risk and severity of infection. The history of influenza vaccination was another critical facilitator of vaccine acceptance attitude, consistent with other studies [7, 58].

### Strengths and Limitations

This systematic review provides updated evidence on the beliefs, attitudes, and acceptance of COVID-19 in both healthcare and non-healthcare workers in the Eastern and Western regions of the world, which can assist in developing region-specific policies to mitigate potential outbreaks of severe COVID-19 variants or other infectious diseases. Nonetheless, this study has several limitations that limit the generalizability of comprehensiveness of the findings. First, we only included peer-reviewed published articles with the potential risk of missing more recent data that may have been available in the grey literature and preprints. Second, all included studies conducted online or through network surveys, which could compromise participant selection, especially with low participant turnout or response rate. Moreover, participation was voluntary in most studies, increasing the risk of selection bias. Third, all included articles were cross-sectional studies, which are typically descriptive, thereby limiting the scope of drawing comprehensive causal inferences. Besides, cross-sectional studies are limited to a particular point in time. Inevitably, people’s attitudes, willingness, and beliefs might have changed over time to influence the acceptance of vaccines.

## Conclusion

Most countries in Eastern and Western regions exhibited low-to-moderate vaccine acceptance rates in the general population and HCWs. Demographic characteristics such as male gender, older age, higher education attainment, and having high socio-economic status strongly related to the positive attitude towards intentions to vaccinate. Other positive predictive factors included self-perceived risks, including fear of contracting the virus, previous history of influenza vaccination, sound knowledge of the vaccine, positive attitude towards the vaccine and its efficacy, trustworthiness of information sources, and mandatory vaccination orders. On the other hand, safety, efficacy, and effectiveness concerns, distrust of the authority or government and information sources, belief in natural immunity, and disinformation were barriers that led to vaccine hesitancy. Therefore, COVID-19 vaccination campaign strategies should be modeled based on regional political, economic, and social contexts for better acceptance.

## Data Availability

All data produced in the present work are contained in the manuscript

## Declarations

### Ethics statement

Since this study utilized publicly available literature, approval from the institutional review board was not obtained.

### Conflicts of interest

The authors declare no conflicts of interest.

### Source of Funding

This study did not receive any research funding.

### Authors’ contribution statement

All authors had access to the data and contributed significantly to writing the manuscript.

## Notes

### Competing Interest Statement

The authors have declared no competing interest.

### Funding Statement

This study did not receive any funding

